# Pathophysiological subtypes of Alzheimer’s disease based on cerebrospinal fluid proteomics

**DOI:** 10.1101/2020.03.25.20043158

**Authors:** Betty M. Tijms, Johan Gobom, Lianne Reus, Iris Jansen, Shengjun Hong, Valerija Dobricic, Fabian Kilpert, Mara ten Kate, Frederik Barkhof, Magda Tsolaki, Frans RJ Verhey, Julius Popp, Pablo Martinez-Lage Alvarez, Rik Vandenberghe, Alberto Lleó, José Luís Molinuevo, Sebastiaan Engelborghs, Lars Bertram, Simon Lovestone, Johannes Streffer, Stephanie Vos, Isabelle Bos, ADNI, Kaj Blennow, Philip Scheltens, Charlotte E. Teunissen, Henrik Zetterberg, Pieter Jelle Visser

**Affiliations:** Alzheimer Center Amsterdam, Department of Neurology, Amsterdam Neuroscience, Vrije Universiteit Amsterdam, Amsterdam UMC, the Netherlands; Clinical Neurochemistry Laboratory, Sahlgrenska University Hospital, Mölndal, Sweden; Department of Psychiatry and Neurochemistry, Institute of Neuroscience and Physiology, Sahlgrenska Academy at the University of Gothenburg, Mölndal, Sweden; Department of Radiology and Nuclear Medicine, Amsterdam Neuroscience, Vrije Universiteit Amsterdam, Amsterdam UMC, Amsterdam, the Netherlands; Institutes of Neurology & Healthcare Engineering, UCL London, London, United Kingdom; 1st Department of Neurology, AHEPA University Hospital, Makedonia, Thessaloniki, Greece; Alzheimer Center Limburg, School for Mental Health and Neuroscience, Maastricht University, Maastricht, the Netherlands; University Hospital Lausanne, Lausanne, Switzerland; Geriatric Psychiatry, Department of Psychiatry, Geneva University Hospital, Geneva, Switzerland; Fundación CITA-Alzhéimer Fundazioa, San Sebastian, Spain; Neurology Service, University Hospitals Leuven, Leuven, Belgium; Laboratory for Cognitive Neurology, Department of Neurosciences, KU Leuven, Leuven, Belgium; IIB-Sant Pau, Hospital de la Santa Creu i Sant Pau, Universitat Autonoma de Barcelona, Barcelona Spain; Barcelonaβeta Brain Research Center (BBRC), Pasqual Maragall Foundation, Barcelona, Spain; Alzheimer’s Disease Unit and Other Cognitive Disorders Unit, Hospital Clinic de Barcelona, Barcelona, Spain; Institute Born-Bunge, Reference Center for Biological Markers of Dementia (BIODEM), Institute Born-Bunge, University of Antwerp, Belgium; Department of Neurology, UZ Brussel and Center for Neurosciences (C4N), Vrije Universiteit Brussel (VUB), Brussels, Belgium; Lübeck Interdisciplinary Platform for Genome Analytics (LIGA), Institutes of Neurogenetics and Cardiogenetics, University of Lübeck, Lübeck, Germany; University of Oxford, Oxford, United Kingdom (currently at Jansen R&D); UCB Biopharma SPRL, Brain-l’Alleud, Belgium; Neurochemistry laboratory, Department of Clinical Chemistry, Amsterdam University Medical Centers (AUMC), Amsterdam Neuroscience, Netherlands; Department of Neurodegenerative Disease, UCL Institute of Neurology, London, United Kingdom; UK Dementia Research Institute at UCL, London, United Kingdom; Department of Neurobiology, Care Sciences and Society, Division of Neurogeriatrics, Karolinska Institutet, Stockholm Sweden

**Keywords:** Alzheimer’s disease, cerebrospinal fluid, proteomics, subtypes

## Abstract

Alzheimer’s disease (AD) is biologically heterogeneous, and detailed understanding of the processes involved in patients is critical for development of treatments. Cerebrospinal fluid (CSF) contains hundreds of proteins, with concentrations reflecting ongoing (patho)physiological processes. This provides the opportunity to study many biological processes at the same time in patients. We studied whether AD biological subtypes can be detected in cerebrospinal fluid (CSF) proteomics using the dual clustering technique non-negative matrix factorization. In two independent cohorts (EMIF-AD MBD and ADNI) we found that 705 (77% of 913 tested) proteins differed between AD (defined as having abnormal amyloid, n=425) and controls (defined as having normal CSF amyloid and tau and intact cognition, n=127). Using these proteins for data-driven clustering, we identified within each cohorts three robust pathophysiological AD subtypes showing 1) hyperplasticity and increased BACE1 levels; 2) innate immune activation; and 3) blood-brain barrier dysfunction with low BACE1 levels. In both cohorts, the majority of individuals was labelled as having subtype 1 (80, 36% in EMIF-AD MBD; 117, 59% in ADNI), 71 (32%) in EMIF-AD MBD and 41 (21%) in ADNI were labelled as subtype 2, 72 (32%) in EMIF-AD MBD and 39 (20%) individuals in ADNI were labelled as subtype 3. Genetic analyses showed that all subtypes had an excess of genetic risk for AD (all p>0.01). Additional pathological comparisons that were available for a subset in ADNI only further showed that subtypes showed similar severity of AD pathology, and did not differ in the frequencies of co-pathologies, providing further support that these differences truly reflect AD heterogeneity. Compared to controls all non-demented AD individuals had increased risk to show clinical progression, and compared to subtype 1, subtype 2 showed faster progression to after correcting for age, sex, level of education and tau levels (HR (95%CI) subtype 2 vs 1 = 2.5 (1.2, 5.1), p = 0.01), and subtype 3 at trend level (HR (95%CI) = 2.1 (1.0, 4.4)). Together, these results demonstrate the value of CSF proteomics to study biological heterogeneity in AD patients, and suggest that subtypes may require tailored therapy.

## Introduction

Alzheimer’s disease (AD) is a neurodegenerative disorder and the most common cause of dementia. The pathological hallmarks are amyloid plaques and tau neurofibrillary tangles in the brain. Biomarkers for amyloid and tau pathology are therefore part of the biological definition of AD (Albert *et al.*, 2011; Dubois *et al.*, 2007; 2014; Jack *et al.*, 2011; 2018; Sperling *et al.*, 2011). The current definition implies that AD is a single disease entity. However, individuals with AD show considerable variability in terms of clinical symptoms, age of onset, disease progression, cortical atrophy patterns, cerebrospinal fluid levels of tau, and other pathological markers (Blennow and Wallin, 1992; Hondius *et al.*, 2016; Iqbal *et al.*, 2005; Lam *et al.*, 2013; Möller *et al.*, 2013; Ossenkoppele *et al.*, 2015; Smits *et al.*, 2015; van der Vlies *et al.*, 2009; Wallin *et al.*, 2010; Whitwell *et al.*, 2012). Part of the heterogeneity in AD is explained by genetic variance, (Ridge *et al.*, 2016) indicating that multiple biological pathways are involved in AD, and these include processes related to amyloid and tau processing, the innate immune system, lipid processing, and synaptic functioning (European Alzheimer’s Disease Initiative (EADI) *et al.*, 2013; Jansen *et al.*, 2019; Kunkle *et al.*, 2019). It is likely that patients will require personalised medicine depending on the molecular processes involved, but at this point there are no tools to identify biological subtypes in AD *in vivo*. Cerebrospinal fluid (CSF) contains many proteins that reflect (patho)physiological processes in the brain, and could provide insight into biological processes involved in AD.

Previous studies examining heterogeneity in AD based on CSF levels of targeted proteins amyloid, tau and p-tau and/or ubiquitin, suggest that at least three subtypes exist, mainly characterised by having low, intermediate or high tau levels (Iqbal *et al.*, 2005; van der Vlies *et al.*, 2009; Wallin *et al.*, 2010). Unbiased or large-scale targeted proteomic CSF analyses have potential to further refine which biological processes become disrupted in AD. So far, AD proteomic studies mostly focussed on finding novel biomarkers by comparing AD individuals with controls (Maarouf *et al.*, 2009; Meyer *et al.*, 2018), and so it remains unclear whether pathophysiological subtypes *within* AD can be discovering with CSF proteomics. Furthermore, if genetic variance in AD risk genes contributes to inter-individual variability in underlying disease mechanisms, it can be hypothesised that these should be detectable already in pre-symptomatic stages of AD.

In this study we used a data-driven dual clustering technique to identify biological subtypes of AD in CSF proteomics in two large independent AD cohorts (i.e., EMIF-AD MBD and ADNI) across the clinical spectrum. We defined AD by the presence of amyloid pathology as indicated by abnormal levels of CSF amyloid β 1-42 (Aβ 1-42), because abnormal Aβ 1-42 CSF shows high concordance with the presence of amyloid and tau pathology upon neuropathological examination (Shaw *et al.*, 2009). In contrast, CSF tau levels show more variability amongst patients, with up to 30% of individuals with pathologically confirmed AD showing normal levels of CSF tau (Shaw *et al.*, 2009). Therefore, we used CSF t-tau and p-tau as independent outcome markers. We further excluded patients that had evidence of known neurodegenerative disorders associated with amyloid aggregation other than AD. We first identified which proteins were associated with AD. Next, we used unsupervised clustering on these proteins to identify biological subtypes of AD. We interpreted AD subtype protein profiles in terms of biological processes through enrichment analyses, and performed post-hoc analyses to characterise AD subtypes in terms of: clinical and biological characteristics known to be associated with AD i.e., established CSF markers (neurogranin, BACE1 activity, neurofilament light, VILIP, YKL-40, sTREM2), *APOE* genotype, AD polygenic risk scores, MRI markers for cortical atrophy, cognitive functioning and decline. Furthermore, we compared subtypes on vascular comorbidity using MRI markers for vascular damage. Finally, we compared subtypes on neuropathological measures that were available for a subset of individuals (ADNI only), and we assessed stability of proteomic subtypes over time for a subset of individuals who had longitudinal proteomics available (ADNI only).

## Methods

### Participant description

We selected individuals with CSF Aβ 1-42, tau, and proteomics data from two independent multicentre AD studies, the European Medical Information Framework for Alzheimer’s disease Multimodal Biomarker Discovery study (EMIF-AD MBD, Bos *et al.*, 2018) and the Alzheimer’s disease Neuroimaging Initiative (ADNI, adni.loni.usc.edu). Both cohorts included individuals with intact cognition, mild cognitive impairment (MCI) or AD-type dementia as determined according to international consensus criteria (McKhann *et al.*, 1984; Mckhann *et al.*, 2011; Petersen *et al.*, 1999; Winblad *et al.*, 2004). Control was defined by intact cognition and normal CSF Aβ 1-42 and tau biomarkers (see next section), and AD pathological change was defined by abnormal CSF Aβ 1-42 (Jack *et al.*, 2018). Both studies excluded patients with any neurologic disease other than suspected AD, such as Parkinson’s disease, dementia with Lewy bodies, frontotemporal dementia, progressive supranuclear palsy, corticobasal syndrome, normal pressure hydrocephalus, and vascular dementia. ADNI started in 2003 as a public-private collaboration under the supervision of Principle Investigator Michael W. Weiner, MD. The primary goal of ADNI is to study whether serial magnetic resonance imaging (MRI), positron emission tomography (PET), other biological markers, and clinical and neuropsychological measures can be combined to measure the progression of mild cognitive impairment (MCI) and early Alzheimer’s disease (AD). Please see www.adni-info.org for the latest information. The institutional review boards of all participating institutions approved the procedures for this study. Written informed consent was obtained from all participants or surrogates.

### Cerebrospinal fluid data

CSF samples were obtained as previously described (Bos *et al.*, 2018; Shaw, *et al.*, 2009; Toledo *et al.*, 2013). CSF Aβ 1-42, t-tau and p-tau levels were measured with INNOTEST ELISAs in EMIF-AD MBD (Bos *et al.*, 2018), and in ADNI with the multiplex xMAP luminex platform (Luminex Corp, Austin, TX) with the INNOBIA AlzBio3 kit (Innogenetics, Ghent, Belgium) at the ADNI Biomarker Core laboratory at the University of Pennsylvania Medical Center. For ADNI biomarker abnormality was defined by Aβ 1-42 levels < 192 pg/ml, and t-tau levels > 93 pg/ml (Shaw *et al.*, 2009). In EMIF-AD MBD cut-offs for p-tau and t-tau were study specific as previously reported (Bos *et al.*, 2018). For Aβ 1-42 cut-offs, the studies in EMIF-AD MBD differed in methodologies used to determine cut-offs, which may lead to bias (Bertens *et al.*, 2017). To minimise such bias across studies, we determined centre specific cut-offs using unbiased Gaussian mixture modelling (see supplementary table 1) (Bertens *et al.*, 2017; De Meyer, 2010; Tijms *et al.*, 2018). Cluster analyses were performed on proteomic data performed using tandem mass tag (TMT) technique with 10+1 plexing in EMIF-AD MBD using high-pH reverse phase HPLC for peptide prefractionation (Batth *et al.*, 2014; Magdalinou *et al.*, 2017; see supplemental methods for more details). For EMIF-AD MBD the median (IQR) analytical CV across included proteins was 5.6 (3.8, 8.0; see supplemental table 2 for protein specific CVs). In ADNI, 4 proteins included were determined with ELISAs, 311 protein fragments determined with Multi Reaction Monitoring (MRM) targeted mass spectroscopy, and 83 proteins measured with Rules Based Medicine (RBM) multiplex. Information on protein assessment and quality control is described at http://adni.loni.usc.edu/data-samples/biospecimen-data/. For ADNI MRM we used the quality controlled finalised ‘Normalized Intensity’ data (Spellman *et al.*, 2015) (please see for detailed explanation of the normalization procedure the “Biomarkers Consortium CSF Proteomics MRM data set” in the “Data Primer” document at adni.loni.ucla.edu). All proteins (EMIF-AD MBD and ADNI) and protein fragment (ADNI) values were first normalised according to mean and standard deviation values of the control group. Next, for ADNI, protein fragments from MRM measurements were combined into a protein score when these correlated with r >.5, and fragments that did not correlate were left out for the present analyses. When the same protein was measured by different platforms in ADNI, values were averaged if they correlated with r>0.5 and else we selected the protein as measured by MRM (one protein was excluded). Only proteins that were observed in 100% of the sample were considered for subsequent analyses, resulting in total 707 proteins in EMIF-AD MBD and 205 proteins in ADNI. A subset of individuals had additional protein measurements available, which we excluded from clustering to use as independent outcomes for subtype interpretation. In ADNI these were Aβ 1-40 and Aβ 1-38 measured with 2D-UPLC tandem mass spectrometry, BACE1 activity, and Elisa measures of neurogranin, neurofilament light, VILIP, YKL40, SNAP25 and sTREM2. In EMIF-AD MBD Elisa measurements were available for Aβ 1-40, Aβ 1-38, neurogranin, neurofilament light, and YKL-40 (Bos *et al.*, 2018). A subset of 70 (29%) ADNI individuals had repeated MRM for 62 proteins (median 5 repeated measures; over median (IQR) of 6 (4.3, 6.7) years for CN and median (IQR) of 4 (3.9, 6.0) years for AD), which we used to study the stability of proteomic subtype. For these analyses, we first standardised the proteins levels according to the baseline mean and SD levels of the control group, and then we constructed proteomic profile scores (PPS) by averaging levels of proteins specific for a subtype.

### Genetic analyses

ADNI samples were genotyped using either the Illumina 2.5-M array (a byproduct of the ADNI whole-genome sequencing sample) or the Illumina OmniQuad array (Saykin *et al.*, 2010). *APOE* genotype was assessed with two SNPs (rs429358, rs7412) that define the epsilon 2, 3, and 4 alleles, using DNA extracted by Cogenics from a 3mL aliquot of EDTA blood. EMIF-AD MBD samples were genotoyped at the USKH site using the Global Screening Array (Illumina, Inc; see (Hong *et al.*, 2019) for more details on imputation preprocessing). In ADNI SNPs were imputed using the 1000 Genomes reference panel, with the use of the Michigan imputation server. Genotype data were quality checked for gender mismatch, relatedness and ancestry. Single nucleotide polymorphisms (SNPs) were excluded prior to data analyses if they had a minor allele frequency less than 2%, deviated significantly from Hardy-Weinberg equilibrium (p<1×10^−6^) in the total sample of founder individuals, or had a call rate of less than 98%. We only used single nucleotide polymorphisms (SNPs) with less than 5% genotype missingness and removed samples with excess heterozygosity rate (>5 SD). After filtering, the genotype data in ADNI included 1,496,949 SNPs and in EMIF-AD MBD 6,706,731 SNPs. To control for population stratification, five principal components were computed on a subset of relatively uncorrelated (r2<0.2) SNPs (PC1-PC5). Polygenic risk scores for AD (PRS) were calculated by adding the sum of each allele weighted by the strength of its association with AD risk using PRSice (Euesden *et al.*, 2014). The strength of these associations was calculated previously by the International Genomics of Alzheimer’s project (IGAP) GWAS (European Alzheimer’s Disease Initiative (EADI) *et al.*, 2013). Clumping was performed prior to calculating PGRS to remove SNPs that are in LD (r^2^<0.1) within a slicing 1M bp window. After clumping we computed fourteen PGRS with varying SNP inclusion threshold (p<10-30 to p<.5). Finally, we constructed specialized PGRS including only SNPs that corresponded to genes part of the GO pathways ‘innate immune response’ and ‘complement activation’ for SNP inclusion thresholds (p<10-30 to p<1). All PGRS were regressed on PC1-PC3.

### Cluster analyses with non-negative matrix factorisation

First, in each cohort we selected proteins for clustering that differed between the control and AD groups at uncorrected p <0.10 using Kruskal-Wallis tests. As protein levels can change non-linearly with levels of neuronal injury and/or disease severity (De Leon *et al.*, 2018; Duits *et al.*, 2018), we repeated analyses stratifying AD individuals on disease stage (i.e., normal cognition, MCI and dementia), and on the presence of abnormal CSF levels of the neuronal injury marker t-tau. Next, we clustered these proteins with non-negative matrix factorisation (NMF). NMF is a dual clustering approach that is based on decomposition of the data by parts, which reduces the dimensionality of data protein expression levels into fewer components which we consider protein profiles (Lee and Seung, 1999), and at the same time this algorithm groups subjects together into subtypes based on how well their protein expression levels match the protein profiles. A strength of NMF compared to correlation-based approaches is that it is able capture non-linear patterns associated with a certain subtype. In order to aid interpretation of the results, we labelled proteins according to which subtype showed the highest average levels. We used the R package NMF for clustering, with the ‘nonsmooth’ option that ensures sparse cluster solutions with enhanced separability (Gaujoux and Seoighe, 2010). The NMF algorithm is stochastic and so subject classification to a subtype can vary from run to run, based on the random initial conditions. We assessed stability of subtype classification over 50 different runs of NMF with the co-phonetic coefficient with values ranging from 0 (i.e., unstable solution) to 1 (i.e., subjects are always classified the same). We tested up to 5 clusters, and the optimal number of clusters was determined as the number of clusters for which: 1. The cophonetic correlation was high; 2. Fit compared to a lower cluster number solution was improved at least 2-fold over a random solution; and 3. Silhouette width of the cluster solution was >.5. Clustering analyses were performed separately for each cohort. We performed pathway enrichment analysis for proteins that were characteristic for each subtype using the online Panther application (Mi *et al.*, 2013). We selected pathways that were most consistently associated with the subtypes for visualisation, and report all observed pathways in the supplementary material. To determine cell type production we used the BRAIN RNASeq database (http://www.brainrnaseq.org, (Y. Zhang *et al.*, 2014).

Proteins were labelled as being specifically produced by a certain cell type when levels were higher than 50% of the total produced across cell types, as non-specific when none of the cell types was higher than 50%, or as not detected when levels were all < 0.2.

### Post-hoc subtype comparisons statistical procedures

We performed the following post-hoc comparisons of subtypes: CSF levels of t-tau, p-tau and other established AD CSF markers that were not included in the cluster analyses to provide further independent interpretation of the cluster solutions, age, gender, disease stage, *APOE* ε4 genotype, AD PGRS, pathological measures, cortical thickness measures from 34 cortical areas as defined by the Desikan-Killiany atlas (averaged over the left and right hemispheres; please see for EMIF-AD MBD (Bos *et al.*, 2018) and for ADNI its website for detailed documentation on variable specific methods: http://adni.loni.cule.edu/), vascular damage (visual ratings in EMIF-AD MBD, and white matter hyperintensity volumes in ADNI), mini-mental state examination (MMSE) scores, level of education, neuropsychological test scores covering the memory (memory immediate and delayed recall scores on the logical memory subscale II of the Wechsler Memory Scale), language (Boston naming test, and animal fluency), visuospatial processing (Clock drawing) and attention/executive domains (digit span, TMT a and TMT b). All continuous variables (except for age, MMSE, and years of education) were standardised according the mean and standard deviation of the control group. Subtype comparisons were performed with general linear models in case of continuous variables with two-sided testing, and with chi square tests for discrete variables. Comparisons for continuous variables were performed without and with adjustment for age and sex, and cognitive measures were additionally adjusted for level of education. We used the R package ‘emmeans’ to obtain estimated marginalised means. ADNI data was downloaded on 30.03.2018. All analyses were performed in R v3.5.1 ‘Feather Spray’.

## Results

We included 127 controls with intact cognition and normal CSF Aβ 1-42 and tau, and 425 individuals with AD across the clinical spectrum (89 (21%) intact cognition, 195 (46%) MCI, and 141 (33%) AD-type dementia). Compared to controls, individuals with AD more often carried an *APOE* ε4 allele, had lower MMSE scores, and more often abnormal CSF p-tau and t-tau in both cohorts (table 1). Other characteristics were similar between groups in both cohorts, except that individuals with AD were older than controls in EMIF-AD MBD. Relative to controls, individuals with AD showed differential CSF levels for 556 of 708 proteins (79%) measured in EMIF-AD MBD and 149 of 205 (73%) proteins measured in ADNI (supplementary table 2). These AD-specific proteins were considered for cluster analyses with NMF within in each cohort.

**Table 1.** Participant description.

| Descriptive | EMIF-AD MBD |  | ADNI |  |
| --- | --- | --- | --- | --- |
|  | Controls (n=82) | Alzheimer's disease (n=228) | Controls (n=45) | Alzheimer's disease (n=197) |
| Cognitive status, n(%): |  |  |  |  |
| Normal cognition | 82 (100%) | 57 (25%) | 45 (100%) | 32 (16%) |
| Mild cognitive impairment | 0 (0%) | 92 (40%)<sup>c</sup> | 0 (0%) | 103 (52%) |
| Dementia | 0 (0%) | 79 (35%) | 0 (0%) | 62 (31%) |
| Age in years, mean (sd) | 61.1 (7)<sup>b</sup> | 68.1 (8)<sup>a, c</sup> | 75.8 (6) | 74.9 (7) |
| Female, n (%) | 47 (57%) | 126 (55%)<sup>c</sup> | 23 (51%) | 81 (41%) |
| Years of education, mean (sd) | 11.9 (3.5)<sup>b</sup> | 11.2 (3.5)<sup>c</sup> | 15.6 (3) | 15.6 (3) |
| MMSE, mean (sd)<sup>1</sup> | 28.6 (1.3)<sup>b</sup> | 25.6 (3.9)<sup>a</sup> | 29.2 (0.6) | 26.1 (2.6)<sup>a</sup> |
| APOE e4, at least one allele (%)<sup>2</sup> | 14 (22%)<sup>b</sup> | 140 (52%) | 4 (8%) | 129 (65%)<sup>a</sup> |
| Hippocampal volume, mean (sd)<sup>3</sup> | 0 (1) | -1.4 (1.5)<sup>a, c</sup> | 0 (1) | -1.7 (1.4)<sup>a</sup> |
| Aβ 1-42 pg/ml, mean (sd) '' | 0 (1) | -2.8 (1.5)<sup>a, c</sup> | 247.5 (29.2) | 139.1 (23.1)<sup>a</sup> |
| T-tau pg/ml, mean (sd) '' | 0 (1) | 4.4 (4.7)<sup>a</sup> | 57.1 (13.1) | 114.3 (54.9)<sup>a</sup> |
| Abnormal t-tau, n (%) ''' | 0 (0%) | 151 (66%)<sup>a</sup> | 0 (0%) | 115 (58%)<sup>a</sup> |
| P-tau pg/ml, mean (sd) '',<sup>4</sup> | 0 (1) | 2.1 (2.5)<sup>a</sup> | 20.3 (9.4) | 39.1 (17.5)<sup>a</sup> |
| Abnormal p-tau, n (%) ''' | 7 (8.5%) | 149 (65%)<sup>a, c</sup> | 9 (20%) | 168 (85%)<sup>a</sup> |

MMSE is mini-mental state examination, APOE is Apolipoprotein E, ICV is total intracranial volume.
' is scaled according to the mean and SD values in controls.
' is based on Luminex in ADNI and scaled for EMIF according to cohort specific controls as previously described (Bos *et al.*, 2018).
''' cut points to define abnormal levels for ADNI: t-tau > 93 pg/ml, p-tau > 21 pg/ml (Shaw *et al.*, 2009), and cohort specific for EMIF-AD MBD as previously described (Bos *et al.*, 2018).
<sup>1</sup> is missing for 1 individual, <sup>2</sup> is missing for 16 individuals, <sup>3</sup> is missing for 159 individuals, <sup>4</sup> is missing for 5 individuals.
Groups were compared with Chi<sup>2</sup> tests or t-test where appropriate.
<sup>a</sup> is p<.01x10<sup>-10</sup> for controls versus AD within cohort.
<sup>b</sup> is p < .05 for across cohort comparisons between controls.
<sup>c</sup> is p < .05 for across cohort comparisons between AD groups.

### Three biological AD subtypes detected in CSF proteomic data

According to our fit criteria, 3 clusters best described the CSF proteomic data in both cohorts (supplementary table 3). Repeating clustering of proteins using a Louvain modularity algorithm on a weighted protein coexpression network also resulted in three protein clusters, which showed good correspondence with the NMF protein clusters of 80% in EMIF-AD MBD and 86% in ADNI (supplementary tables 5a and 5b). A 3D plot of subject loadings on clusters revealed in EMIF-AD MBD a subset of 5 individuals with extreme loadings (supplementary figure 1). These individuals did not show differences with other AD individuals in terms of sample characteristics (supplementary table 4). To avoid potential overfitting, we repeated cluster analyses excluding these individuals, and a three-cluster solution remained most optimal. We next labelled individuals according to the subtype they scored highest on (figure 1a). In both cohorts, the majority of individuals was labelled as having subtype 1 (80, 36% in EMIF-AD MBD; 117, 59% in ADNI), 71 (32%) in EMIF-AD MBD and 41 (21%) in ADNI were labelled as subtype 2, 72 (32%) in EMIF-AD MBD and 39 (20%) individuals in ADNI were labelled as subtype 3. A subset of 92 proteins was measured in both EMIF-AD MBD and ADNI, which showed consistent subtype differences in levels for 84-98% of proteins across the cohorts, further supporting that subtype definitions are robust (supplementary figure 2). Individuals with subtype 1 had compared to controls significantly higher levels for the majority of proteins in both cohorts (EMIF-AD MBD: 309, 56%; ADNI: 92, 65%; p values ranging between .02×10^−21^ and .049; figure 1b; supplementary tables 5a and 5b). The predominant cell types producing these proteins were neurons and astrocytes (figure 1c; supplementary table 6). GO pathway analyses for proteins increased in subtype 1 showed enrichment for processes MAPK/ERK cascade, synaptic structure and function, axonal development, and glucose metabolism, suggesting that subtype 1 shows neuronal hyperplasticity (figure 1d; supplementary table 7). Subtype 2 also showed mostly protein higher levels than controls (EMIF-AD MBD: 202, 36%; ADNI: 31, 21%; p values ranging between .01×10^−16^ and .049). The predominant cell types producing these proteins were oligodendrocytes, neurons and astrocytes. GO pathway analyses for proteins specifically increased in subtype 2 showed enrichment for innate immune response, complement activation, extracellular matrix organisation and oligodendrocyte development, hence these individuals may be characterised as having innate immune activation. Compared to controls, subtype 3 individuals showed mostly decreased proteins (415, 75% in EMIF-AD MBD; 120, 81% in ADNI; p values ranging between .02×10^−22^ and .049) that mirrored the increases observed in subtype 1, which suggests that type 3 has neuronal hypo-plasticity. Another group of proteins was specifically increased in Subtype 3 compared to controls (76, 14% in EMIF-AD MBD; 6, 4%; in ADNI), including albumin and immunoglobulin proteins, of which higher CSF levels have been reported with blood-brain barrier dysfunction (Dayon *et al.*, 2019). GO pathway analyses for proteins specifically increased in subtype 3 showed enrichment for acute inflammation, b-cell activation, blood coagulation-related processes, lipid processing, and lipoprotein clearance, which together suggest that this subtype may be characterised as having blood-brain barrier dysfunction. Subtype 3 also showed enrichment for complement activation, but for a different group of proteins than observed in subtype 2: C6, C8A, C8B and C9, which are part of the terminal pathway of the complement system (Orsini *et al.*, 2014; Veerhuis *et al.*, 2011; supplementary figure 3). Longitudinal proteomics was available for a subset in ADNI (n=70 (29%), including 23 controls and 47 AD: 31 with subtype 1; 9 with subtype 2; 7 with subtype 3), including only proteins associated with subtype 1 and 2. Proteomic profile scores (PPS) that summarised levels of proteins that were associated with either subtype 1 (52 proteins) or 2 (12 proteins) in the discovery dataset remained stable over time in all subtypes, as none of the slopes differed from 0 (all subtypes p >0.10, supplementary table 8). This suggests that subtype definitions remained stable over time.

**Figure 1.**
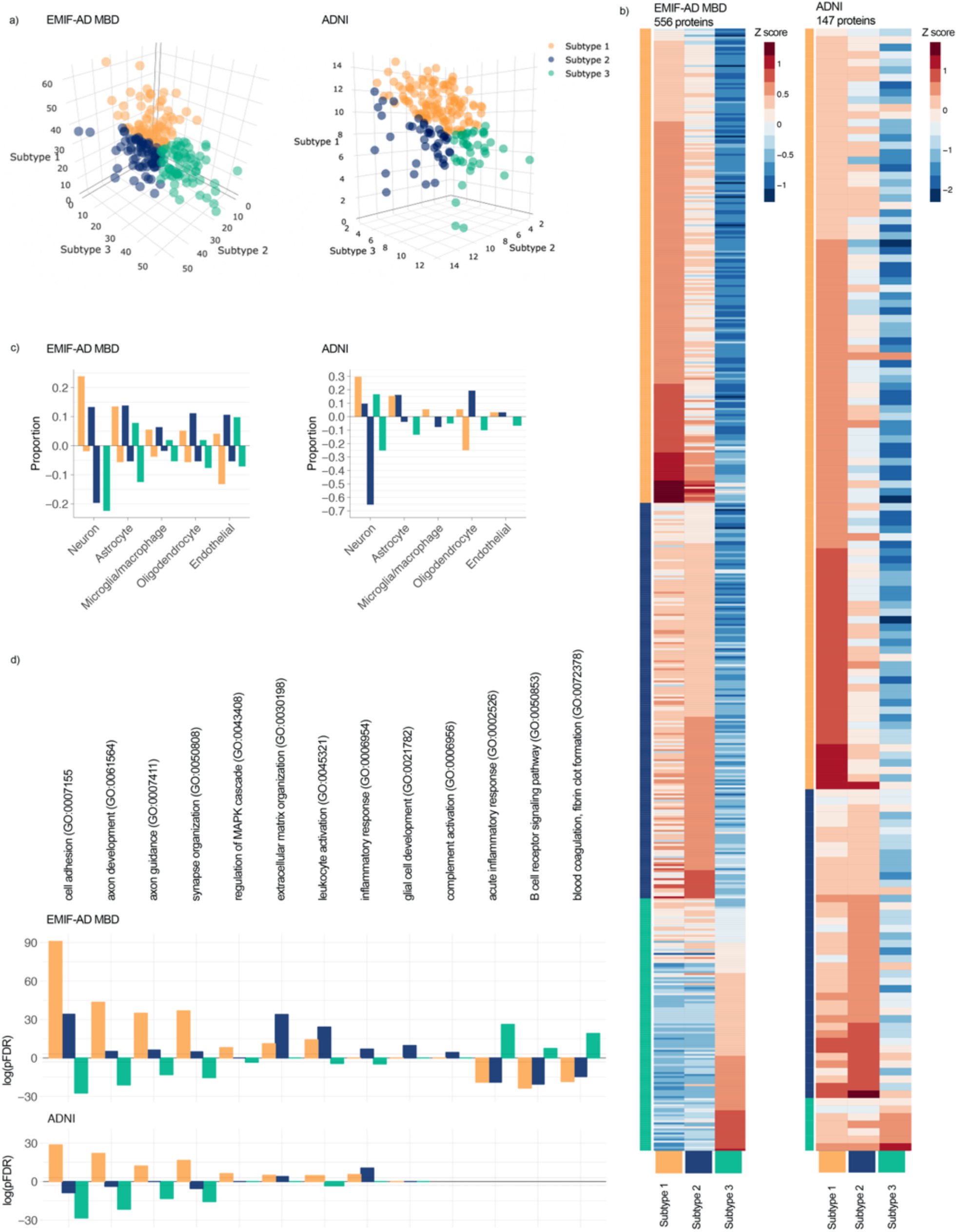
a) Subject loadings on subtype scores (orange is subtype 1, hyperplasticity; blue is subtype 2, innate immune activation; green is subtype 3 blood-brain-barrier dysfunction) for EMIF-AD MBD (left) and ADNI (right). Each dot shows how an individual matches all three proteomic subtypes at the same time. E.g., the right most green dot is a person who shows very high loading on the subtype 3 axis, and very low loadings on subtype 1 and 2 axes. b) Heatmap of subtype average Z-scores (according to the mean and SD of controls), labels not shown, see for list of proteins supplementary table 5. c) Proportion of cell type production for protein levels higher (positive proportions) or lower than controls (negative proportions). d) Selected subset of GO pathways that show subtype specific enrichment with log(pFDR) positive values for proteins with higher levels than controls, and negative values for proteins with lower levels than controls (see supplementary table 7 for complete list of enriched pathways).

### Genetic comparisons of subtypes

Subtypes showed similar proportions of *APOE* ε4 carriers in both cohorts (figure 2a; all p>0.05; supplementary table 9). Relative to controls, all subtypes had an excess of AD genetic risk (figure 2b, supplementary table 10; p values ranging between .02×10^−13^ and .004). For SNP inclusion thresholds .1 to .5, Subtype 2 individuals showed higher AD PRS than subtype 1 and 3, but these associations lost significance after adjusting for age and sex. Because Subtype 2 individuals were associated with innate immune response, which has been previously associated with top AD risk SNPs, we compared subtypes on PRS for innate immune response and complement activation, and found for the majority of SNP inclusion thresholds the highest scores for subtype 2 (figure 2b; supplementary table 10; p values compared to controls ranging between .02×10^−7^ and .045; p values compared to the other subtypes ranging between .004 and .045). These effects remained largely unchanged after adjusting for age and sex.

**Figure 2.**
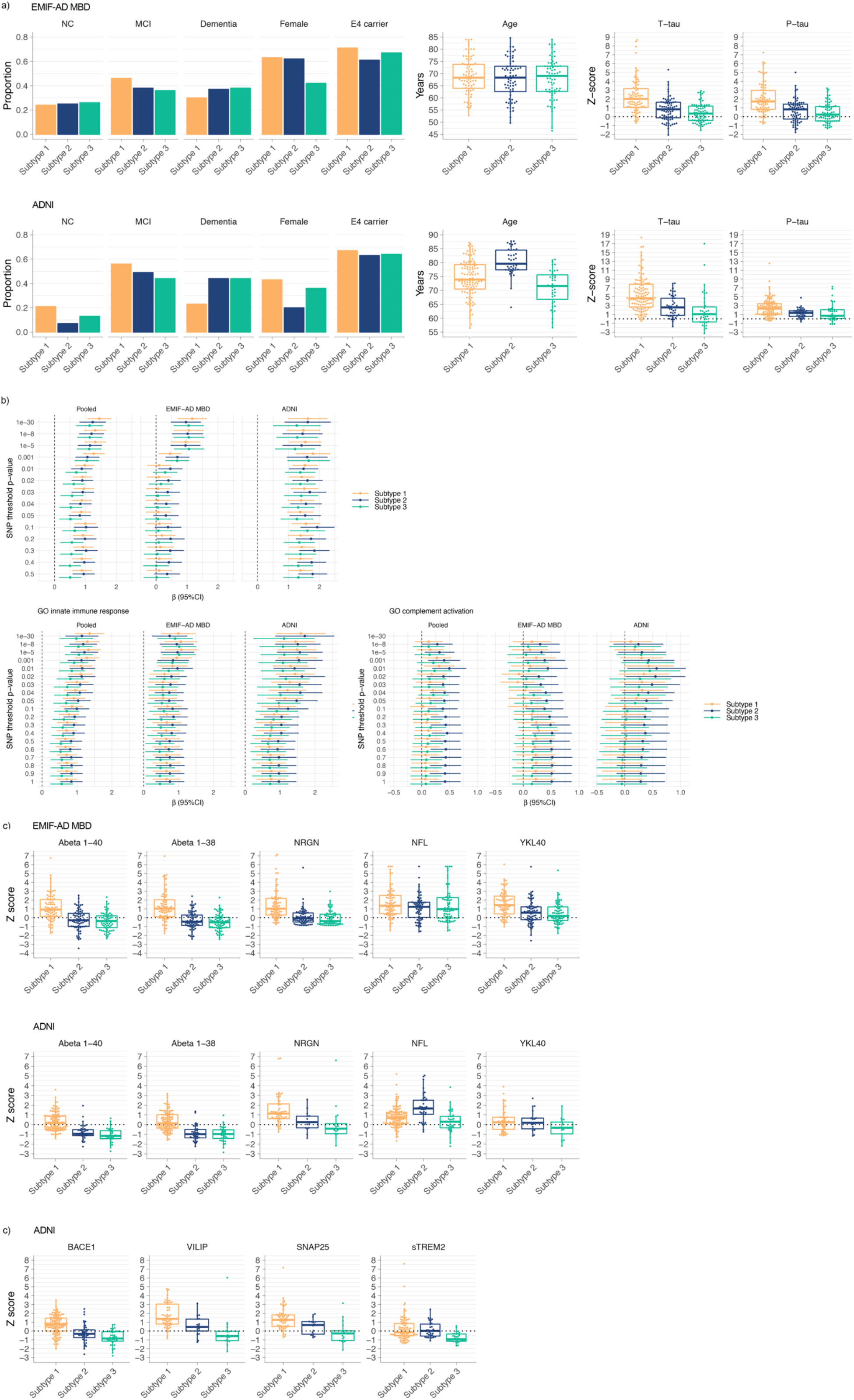
Subtype comparisons on: a) proportions of disease stage, females, and *APOE* e4 carriers, and distributions of age, t-tau and p-tau levels for EMIF-AD MBD (top row) and ADNI (bottom row); c) CSF markers not included in clustering for EMIF-AD MBD (top row) and ADNI (bottom row); b) Effect sizes (95%CI) as compared to the control group for Polygenic Risk Scores (PRS) for AD including all SNPs for various p value inclusion thresholds, and two similar figures with AD PRS restricted to genes involved in GO innate immune response and GO complement activation. Plots are shown for the pooled sample, and for the cohorts separately. Dotted vertical line indicates scores for the control group. c) Additional CSF biomarkers that were available in ADNI for a subset of individuals. See supplementary tables 8 and 15 for test statistics of all comparisons.

### Other biological and clinical subtype characterisation

We next compared subtypes on clinical characteristics and established AD CSF markers. In EMIF-AD MBD subtypes had comparable age and proportions of disease stages and sex. In ADNI, individuals with subtype 1 (hyperplasticity) less often had dementia (compared to subtype 2 p = .02; compared to subtype 3 p = .02), and individuals with subtype 2 (innate immune activation) were older and more often male. In both cohorts, t-tau and p-tau CSF levels were highest and most often abnormal in the subtype 1(hyperplasticity; figures 2a; supplementary table 9), intermediate for subtype 2 (innate immune activation), and the lowest and most often normal in subtype 3 (blood-brain barrier dysfunction). Other neuronal injury markers such as neurogranin (both cohorts), VILIP and SNAP25 (ADNI only) were consistently highest in subtype 1 (hyperplasticity), and lowest in subtype 3 (blood-brain barrier dysfunction) (figures 2c-d). NEFL levels were comparable across subtypes in EMIF-AD MBD, but were increased in subtype 2 (innate immune activation) in ADNI, which remained after additional correction for age and sex. Subtype 1 (hyperplasticity) further showed higher levels of proteins associated with amyloid precursor protein (APP) processing (i.e., higher levels of Aβ 1-40 and Aβ 1-38 in both cohorts, and higher levels of BACE1 activity in ADNI). Subtype 3 (blood-brain barrier dysfunction) showed the lowest concentrations for those markers. Both subtype 1 (hyperplasticity) and 2 (innate immune activation) showed higher levels of inflammation markers YKL40 and sTREM2 (ADNI only) than subtype 3 (blood-brain barrier dysfunction). Since some of these markers can increase with disease severity, we repeated subtype comparisons stratified for disease stage (intact cognition, MCI and dementia). Results showed largely similar subtype profiles (all p_interaction_ >.05; supplementary figures 4, 5).

### Atrophy, vascular damage, cognitive profiles and pathological comparisons

Atrophy relative to controls was most pronounced in the hippocampus, medial and lateral temporal cortex and the precuneus for all subtypes (figure 3a; supplementary table 11; supplementary figure 6). Compared to subtype 1 (hyperplasticity), individuals with subtype 2 (innate immune activation) and 3 (blood-brain barrier dysfunction) subtype showed more atrophy in the posterior cingulate in both cohorts (figure 3b). In ADNI, subtype 2 (innate immune activation) showed more atrophy than subtype 1 (hyperplasticity) in the inferior temporal gyrus, insula, isthmus cingulate, rostral middle frontal and temporal pole. Visual ratings for vascular damage on MRI in EMIF-AD MBD showed that subtype 3 (blood-brain barrier dysfunction) more often had a lacunar infarct (n=10, 22%) than subtype 2 (1, 2%; p=.003) and subtype 1 (4, 7.5%; p=.04). No differences between subtypes were observed in white matter intensity load (Fazekas score of 3), or the presence of more than 1 microbleed (supplementary table 8). In ADNI white matter hyperintensity volumes were larger in subtype 3 (blood-brain barrier dysfunction; 1.2 ± 2.7 cm^3^) and subtype 2 (innate immune activation; 1.3 ± 1.4 cm^3^) compared to subtype 1 (hyperplasticity; 0.85 ± 3.0 cm^3^; 1vs2 p = .0004; 1vs3 p =.01; 2vs3 p =.44). Subtypes showed largely similar scores on cognitive tests (figure 3c; supplementary table 11). Repeating analyses stratified on disease stage, showed that individuals with subtype 3 in the dementia stage scored worse on the TMTa than the other two subtypes (p_interaction_ = .004; both 1vs3 and 2vs3 p < .001; supplementary table 12; supplementary figure 7). Worsening over time on CDRsob was steeper for subtype 2 compared to subtype 1 in MCI (p=0.01; figure 3e, f; supplementary table 13), and for subtype 3 compared to subtype 1 in dementia (p=.02). Compared to subtype 1, individuals without dementia and subtype 2 showed increased risk to progression to dementia, also after correcting for age, sex, level of education and tau levels (HR (95%CI) subtype 2 vs 1 = 2.5 (1.2, 5.1), p = 0.01), and subtype 3 at trend level (HR = 2.1 (1.0, 4.4), p=.06; figure 3g; supplementary table 14). For a subset of 20 (10%) ADNI individuals with neuropathological information, we found similar pathological scores for amyloid and tau for subtypes, and they showed similar frequencies of occurring co-pathologies, such as Lewy body pathology, TDP-43 and hippocampal sclerosis (supplementary table 15).

**Figure 3.**
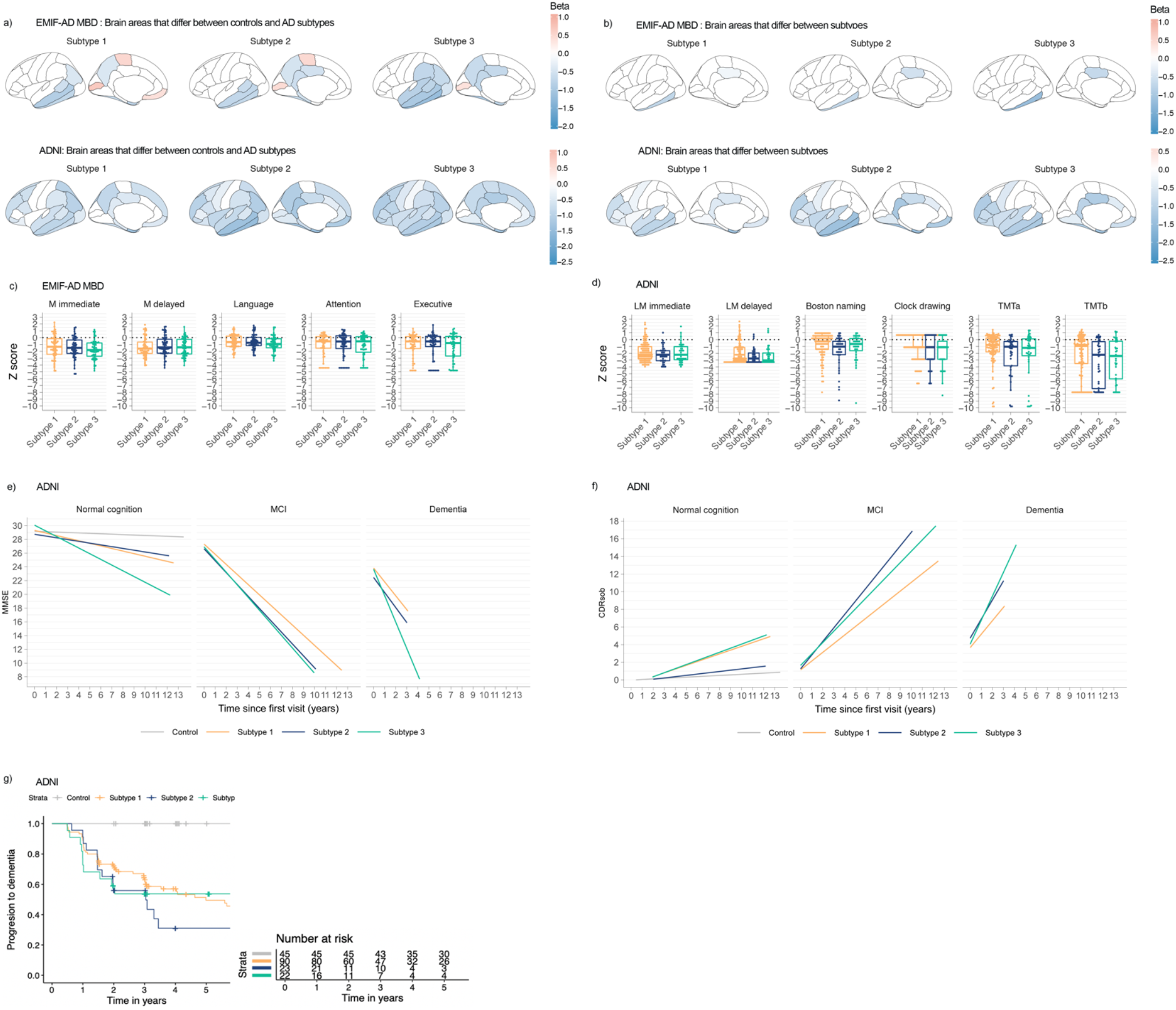
Cortical thickness comparisons between subtypes against controls, a) shows all brain areas were differences against controls were observed, b) shows brain areas with a significant main effect for subtype; all beta values reflect volumetric differences of subtypes against controls. C and d) Comparisons of cognitive profiles between subtypes. Changes over time on MMSE (e), and CDRsob (f) in ADNI only. g) Cumulative progression to dementia curves for subtypes, in ADNI only. All cortical thickness and neuropsychological test values are standardized according to the mean and standard deviation values of the controls. See supplementary table 9, 10, 18 and 19 for test statistics of all comparisons.

## Discussion

Understanding biological heterogeneity in patients with AD is critical for treatment development. We proteomically defined three AD pathophysiological subtypes that were associated with distinct biological processes, i.e., hyperplasticity, innate immune activation and blood-brain barrier dysfunction, and these subtypes were robustly observed in two large independent cohorts. These biological subtypes of AD showed pronounced differences in levels of proteins associated with processes known to be deregulated in AD including APP processing, neuronal injury, and inflammation. All subtype specific alterations in CSF protein levels could both be in- or decreased for subsets of proteins, indicating that differences cannot be explained by trivial a-specific changes in CSF composition. All subtypes had an excess genetic risk for AD, and pathological measures did not show a difference in the presence of comorbidities, providing further support that these differences reflect heterogeneity *within* AD. A particularly novel finding is the observation of the blood-brain barrier dysfunction subtype that showed mostly abnormally low concentrations of proteins associated with APP processing, as well as t-tau and p-tau levels. Together, these results demonstrate the potential for CSF proteomics to identify which biological processes are disrupted in individual persons with AD, and suggest that individuals might require specific treatments depending on their subtype.

Previous studies that clustered targeted proteins amyloid, t-tau, p-tau and/or ubiquitin CSF levels suggested three to five AD subtypes that were characterised by having low, intermediate and high tau values (Iqbal *et al.*, 2005; van der Vlies *et al.*, 2009; Wallin *et al.*, 2010), and our proteomically defined subtypes show a similar distinction in tau levels. We further show with our large-scale proteomic analyses which biological processes might underlie interindividual differences in tau levels. AD individuals with the hyperplasticity subtype showed high levels for the majority of proteins, which were enriched for regulation of MAPK/ERK cascade, glucose metabolism, synaptic structure and function, and axonal development, all processes important for synaptic plasticity. This hyperplasticity subtype also showed higher levels of markers presumed to reflect neuronal injury, i.e., t-tau, p-tau, neurogranin, VILIP, and SNAP25, which could reflect more severe neuronal damage (Brinkmalm *et al.*, 2014; Fagan and Perrin, 2012). However, this is unlikely, because these proteins were already increased in individuals with AD and normal cognition, when atrophy was less severe compared to the other subtypes. An alternative explanation could be increased synaptic activity as this can lead to increased tau (Pooler *et al.*, 2013; Yamada *et al.*, 2014) and amyloid secretion (Bero *et al.*, 2011; Cirrito *et al.*, 2005). Hyperactive neurons have been reported in AD with concurrent increased tau and amyloid levels (Palop and Mucke, 2016; Roberson *et al.*, 2011). Aberrant increases in neuronal activation can be caused by amyloid oligomers, which disrupt the balance of excitation and inhibition of neuronal circuits (Palop and Mucke, 2016).

The second subtype had a proteomic profile that also indicated upregulation of plasticity-related processes like subtype 1, but less pronounced. Subtype 2 further showed higher levels of proteins involved in the classical complement pathway, extracellular matrix organization and oligodendrocyte development. Oligodendrocytes are important for axonal myelination. In ADNI, subtype 2 showed increased white matter hyperintensity volume, and in both EMIF-AD MBD and ADNI NEFL was increased, suggesting axonal damage in this subtype. Subtype 2 specifically showed higher levels of MMP2, which plays a role in degradation of the extracellular matrix, is produced by microglia and oligodendrocytes, and can lead to axonal damage (Diaz-Sanchez *et al.*, 2006). Furthermore, individuals with subtype 2 showed high levels of complement proteins C1q B chain and C4a, and in EMIF-AD MBD also C1q A chain, C1q C chain, C1s, and C1r, which are early components of the classical complement pathway (Orsini *et al.*, 2014; Veerhuis *et al.*, 2011). AD polygenic risk scores restricted to genes involved in innate immune response and complement activation were mostly higher in subtype 2, suggesting that these CSF proteomic alterations reflect genetic effects. Higher concentrations of C1q and C4 in AD brains have been reported in pathological studies (Veerhuis *et al.*, 2011); (Dejanovic *et al.*, 2018), and so higher concentrations of C4a might indicate complement activation in this subtype. Amyloid beta fibrils are known to activate the complement pathway by binding to the C1q complex (Rogers *et al.*, 1992; Webster *et al.*, 2002). Complement activation might also play a role in neuronal injury in AD, because complement proteins can accumulate at synapses and tag these for phagocytosis by activated microglia (Orsini *et al.*, 2014; Pooler *et al.*, 2013; Yamada *et al.*, 2014). Knocking out C1q in APP transgenic mice or blocking C1q in Tau-P301S transgenic mice attenuates both complement activation and neuronal injury (Orsini *et al.*, 2014; Pooler *et al.*, 2013; Yamada *et al.*, 2014), possibly by preventing inappropriate microglia activation. Together, the biological processes specific for subtype 2 seem associated with activated microglia. Alternatively, these processes may be associated with activation or dysregulation of astrocytes, because microglia secreting C1q can induce so-called ‘A1 reactive’ astrocytes that lose the ability to facilitate plasticity processes that promote cell survival and accelerate death of neurons and oligodendrocytes (Liddelow *et al.*, 2017).

Subtype 3 had low and more often normal t-tau and p-tau CSF levels compared to the other subtypes, together with abnormally low levels for the majority of other proteins. The generally low levels of t- and p-tau raise the question as to whether these individuals have AD (Jack *et al.*, 2018). Our results provide strong support that in fact these individuals do have AD, because: 1. these individuals had abnormal amyloid levels; 2. the most severe atrophy included typical AD regions such as the medial temporal lobe; 3. non-demented individuals with subtype 3 showed increased risk for clinical progression; and 4. they have an excess of genetic risk for AD. The percentage of individuals classified as having subtype 3 is in line with previous studies reporting in pathologically confirmed AD, which show that up to 30% of individuals can have normal CSF tau levels (Shaw *et al.*, 2009). Thus, CSF tau levels may reflect other processes in addition to neurofibrillary tau tangles, and conversely, normal levels do not exclude underlying tau pathology. Given the relationship of tau levels and neuronal activity discussed above, low tau levels in this subtype may reflect hypo-plasticity. Alternatively, low levels of tau suggest less neuronal injury. However, this explanation seems implausible, because subtype 3 had widespread atrophy and high levels of the axonal damage marker NEFL. Other proteins that were increased in Subtype 3 have previously been reported to correlate with the CSF/plasma albumin ratio, which is a marker for blood-brain barrier integrity (Dayon *et al.*, 2019), and this indicates that subtype 3 may have blood-brain barrier dysfunction (Sagare *et al.*, 2012; Sweeney *et al.*, 2018; Yu YamazakiTakahisa Kanekiyo, 2017). Blood-brain barrier dysfunction disrupts glucose metabolism, which can impair neuronal activity and plasticity processes (Sweeney *et al.*, 2018; Yamazaki and Kanekiyo, 2017). This would explain why proteins involved in synaptic structure and function were decreased in subtype 3, and suggest that these individuals have hypo-plasticity. Another subset of proteins specifically increased in subtype 3 was enriched for lipid processing, clearance and regulation, and these included apoC1. ApoC1 is produced by astrocytes (Abildayeva *et al.*, 2008; Petit-Turcotte *et al.*, 2001), can inhibit receptor mediated clearance of lipoproteins containing *APOE* (Sehayek and Eisenberg, 1991; Shachter, 2001), and has been observed in amyloid plaques (Abildayeva *et al.*, 2008). This suggests that vascular factors might play a role in amyloid pathogenesis, possibly contributing to reduced clearance of aggregated amyloid. Alternatively, amyloid might aggregate in the vasculature which could lead to blood-brain barrier dysfunction.

The proteomic subtypes we discovered could have implications for treatment: Subtype 1 showed the highest levels of BACE1 activity and products of amyloid metabolism (Aβ 1-40 and Aβ 1-38) and so it can be hypothesised that particularly this subtype will benefit from treatments that target APP processing, such as BACE1 inhibitors, whereas this type of treatment may be harmful for individuals with subtype 3 that showed decreased levels of BACE1 activation. Individuals with subtype 2 may potentially benefit from therapeutic strategies that target microglia and astrocyte activation. Subtype 3 may benefit from therapies that protect the vasculature. Future research should further study treatment effects on CSF proteomic profiles, and whether effects are subtype dependent.

A potential limitation of this study is that proteins specifically increased in subtype 3 were mostly observed in EMIF-AD MBD, because that study used an untargeted approach, whereas those proteins were not measured in ADNI, because that study selected a limited number of proteins with brain enriched expression patterns. Still, also in ADNI subtype 3 showed a hypo-plasticity response, highly similar to that of subtype 3 in EMIF-AD MBD, suggesting that they share common pathophysiological processes. Also, although many of the proteins specifically increased in subtype 3 were previously reported to be correlated to blood-brain barrier function (Dayon *et al.*, 2019), future analyses should further verify this by measuring both CSF and plasma albumin. Furthermore, the EMIF-AD MBD and ADNI cohorts, are respectively a clinical multicentre and a research multicentre study, and differed in their baseline characteristics, most notably ADNI participants being on average 10 years older. However, it is unlikely cohort specific- and age effects explain the subtypes, since the subtypes showed highly similar proteomic profiles across cohorts. Furthermore, MRI scans in EMIF-AD MBD were acquired in clinical routine, in contrast to the harmonized scanning protocol in ADNI. This may made it more difficult to detect differences in atrophy patterns amongst subtypes in combination with relatively small sample sizes (Kate *et al.*, 2018; Scheltens *et al.*, 2017; X. Zhang *et al.*, 2016). Another point of consideration is that with older age co-pathology often occurs, which may influence proteomic subtype definitions (Beach *et al.*, 2012). Our analyses in a subset of ADNI showed that subtypes did not differ in the occurrence of co-pathology, suggesting that it is unlikely that this has driven the subtypes. Still, those analyses need replication in larger samples. Our subtypes were defined by their proteomic profiles, and at this point single markers from our analyses should not be used in practice until thoroughly validated, which we aim to pursue in future studies. A strength of our study is that we were able to replicate the biological subtypes that we detected with CSF proteomics in two independent cohorts, and even though different methods were used to measure proteins we observed similar processes to be involved in AD, supporting the robustness of our findings.

In conclusion, we have identified a hyperplasticity, innate immune activation and a blood-brain barrier dysfunction subtype in AD using CSF proteomics. The most important implication of our results is that currently existing- and even failed-treatments might be beneficial for specific subtypes, and that CSF proteomics may serve as a stratification tool to further investigate this.

## Data Availability

Data used in preparation of this article were obtained from the Alzheimer’s Disease Neuroimaging Initiative (ADNI) database (adni.loni.usc.edu). EMIF-AD MBD data are available for research purposes upon request.

## Acknowledgements

This work has been supported by ZonMW Memorabel grant programme #73305056 (BMT) and #733050824 (BMT and PJV), the Swedish Research Council (#2018-02532, HZ), the European Research Council (#681712, HZ) and Swedish State Support for Clinical Research (#ALFGBG-720931, HZ), and the Innovative Medicines Initiative Joint Undertaking under EMIF grant agreement #115372 (PJV, HZ). Statistical analyses were performed at the VUmc Alzheimer Center that is part of the neurodegeneration research program of the Neuroscience Campus Amsterdam. EMIF-AD MBD proteomic analyses were performed at the Department of Psychiatry and Neurochemistry, the Sahlgrenska Academy at the University of Gothenburg, Sweden. The VUmc Alzheimer Center is supported by Stichting Alzheimer Nederland and Stichting VUmc fonds. HZ is a Wallenberg Academy Fellow. FB is supported by the NIHR biomedical research centre at UCLH. The Leuven cohort was funded by Stichting Alzheimer Onderzoek (#11020, #15005, #13007) and the Vlaamse Impulsfinanciering voor Netwerken voor Dementie-onderzoek (IWT #135043).

Data was used for this project of which collection and sharing was funded by the Alzheimer’s Disease Neuroimaging Initiative (ADNI) (National Institutes of Health Grant U01 AG024904) and DOD ADNI (Department of Defense award number W81XWH-12-2-0012). ADNI is funded by the National Institute on Aging, the National Institute of Biomedical Imaging and Bioengineering, and through generous contributions from the following: AbbVie, Alzheimer’s Association; Alzheimer’s Drug Discovery Foundation; Araclon Biotech; BioClinica, Inc.; Biogen; Bristol-Myers Squibb Company; CereSpir, Inc.; Cogstate; Eisai Inc.; Elan Pharmaceuticals, Inc.; Eli Lilly and Company; EuroImmun; F. Hoffmann-La Roche Ltd and its affiliated company Genentech, Inc.; Fujirebio; GE Healthcare; IXICO Ltd.; Janssen Alzheimer Immunotherapy Research & Development, LLC.; Johnson & Johnson Pharmaceutical Research & Development LLC.; Lumosity; Lundbeck; Merck & Co., Inc.; Meso Scale Diagnostics, LLC.; NeuroRx Research; Neurotrack Technologies; Novartis Pharmaceuticals Corporation; Pfizer Inc.; Piramal Imaging; Servier; Takeda Pharmaceutical Company; and Transition Therapeutics. The Canadian Institutes of Health Research is providing funds to support ADNI clinical sites in Canada. Private sector contributions are facilitated by the Foundation for the National Institutes of Health (www.fnih.org). The grantee organization is the Northern California Institute for Research and Education, and the study is coordinated by the Alzheimer’s Therapeutic Research Institute at the University of Southern California. ADNI data are disseminated by the Laboratory for Neuro Imaging at the University of Southern California.

## Author Contributions

Study design: BMT, PJV.

Design proteomics EMIF-AD MBD: JG, HZ.

Design statistical analyses: BMT, PJV.

Data acquisition, processing, technical: JG, LR, IJ, SH, VD, FK, MtK, FB, MT, FRJV, JP, PMLA, RV, AL, JLM, SE, LB,

SL, JS, SV, IB, KB, PS, CET, HZ, PJV.

Manuscript drafting: BMT, PJV. Manuscript revising: all authors.

